# TELF: An End-to-End Temporal Encoder with Late Fusion for Interpretable Disease Risk Prediction from Longitudinal Real-World Data

**DOI:** 10.64898/2026.04.04.26350180

**Authors:** Yunhao Liu, Zhiwei Zhang

## Abstract

Deep learning models utilizing longitudinal healthcare data have significantly advanced epidemiological research. However, contemporary transformer-based models increasingly rely on computationally intensive pre-training steps that entail processing massive real-world datasets with cost-prohibitive hardware. We introduce the <u>T</u>emporal <u>E</u>ncoder with <u>L</u>ate <u>F</u>usion (TELF), a lightweight end-to-end predictive model featuring an encoder-only architecture for processing medical codes, followed by post-encoder concatenation with demographic variables. TELF learns code embeddings on-the-fly, thereby bypassing the resource-intensive pre-training bottleneck. Furthermore, its late-fusion design preserves the integrity of the temporal attention mechanism before integrating static demographic predictors. We evaluated TELF using an administrative claims database across three distinct cohorts: pancreatic cancer (n=53,661), type 2 diabetes (n=78,756), and heart failure (n=72,540). TELF consistently outperformed traditional machine learning baselines, including XGBoost, LightGBM, and logistic regression. Specifically, TELF achieved AUCs of 0.9150, 0.8199, and 0.8721 for pancreatic cancer, type 2 diabetes, and heart failure, respectively, compared with 0.9044, 0.7908, and 0.8535 for XGBoost and 0.9014, 0.7800, and 0.8466 for logistic regression. Beyond predictive superiority, TELF’s isolated temporal attention mechanism enables population-level motif mining. By extracting high-attention temporal sequences, we mapped aggregated patient journey pathways, revealing interpretable clinical trajectories preceding disease onset. Collectively, these results demonstrate that TELF provides a resource-efficient and accessible framework for advanced temporal modeling in clinical and epidemiological research.

## INTRODUCTION

The recent evolution of deep learning (DL) in disease prediction has profoundly benefited from the synergy between large-scale longitudinal healthcare data and state-of-the-art transformer model [1] and its self-supervised derivative bidirectional encoder representations from transformers (BERT) [2]. Recognizing the inherently chronological nature of patient journeys, researchers rapidly adapted these self-attention-based architectures to clinical risk prediction. Foundational two-step models, such as BEHRT, ClinicalBERT, and Med-BERT [3–6], demonstrated significant performance gains over traditional machine learning methods by treating medical codes as elements of a temporal sequence. These models predominantly operate on a two-step paradigm [7]. In the first step, massive corpora of real-world data are processed via self-supervised pre-training to learn universal vector representations (embeddings) of medical codes. This phase often necessitates access to specialized high-performance computing clusters. In the second step, the pre-trained embeddings are transferred and fine-tuned on a smaller, condition-specific cohort to execute a targeted downstream prediction task.

There has been a trend toward increasingly complex architectures for modeling temporal medical code sequences. Recent models, such as TransformEHR [8] and EHRFormer [9], employ full encoder-decoder structures rather than the encoder-only designs of their BERT-based predecessors. Despite these architectural differences, a critical limitation unites these contemporary approaches: they rely heavily on computationally intensive self-supervised pre-training. Processing large EHR corpora to learn code representations requires cost-prohibitive computing hardware and prolonged training times (Table 1). Consequently, these models remain largely inaccessible to many clinical researchers, concentrating advanced predictive capabilities within a small number of well-resourced institutions rather than enabling broader adoption across the research community.

**Table 1.** Computing hardware and training times of representative resource-intensive foundation models.

|  | BioBERT | Med-BERT | GatorTron | TransformEHR |
| --- | --- | --- | --- | --- |
| Input modality | Unstructured Biomedical Text | ICD-9/ICD-10 | Unstructured Clinical Notes | ICD-10CM and demographics |
| Pre-training data source | PubMed and PMC | Cerner Health Facts EHR | UF Health IDR and supplements | US Veterans Health (VHA) |
| Number of pre-training patients | N/A (18 billion words) | 20 million | 2.5 million | 6.5 million |
| Model size (parameters) | 110 million | ~17 million | Up to 8.9 billion | Matched to BERT (~110 million) |
| Training resources | 8 Tesla V100 32GB GPUs (23 days) | A Tesla V100 32GB GPU (a week) | 992 A100 80GB GPUs (~6 days) | 4 Tesla P40 22GB GPUs (6 days) |

Furthermore, many existing models are developed specifically for electronic health record (EHR) data and are often named to reflect this focus [3, 8, 9]. While EHR data provides rich clinical detail, they frequently suffer from enrollment ambiguity [10], forcing researchers to rely on encounter-based proxies to approximate enrollment periods [11]. In contrast, administrative claims databases offer the distinct advantage of verifiable continuous enrollment, ensuring the uninterrupted and high-quality capture of medical code sequences over a defined baseline period.

Additionally, when processing these sequences, existing transformer-based models often employ early fusion to integrate static demographic variables (such as sex and race) as embeddings [3, 8, 12–14]. This approach implies that the representation of a medical code varies according to demographic characteristics, an assumption that is rarely justified clinically and may confound the interpretation of the temporal attention mechanism. In contrast, late fusion architectures treat demographic variables as risk modifiers, incorporating them only after the temporal sequence representation has been learned [15].

To address these computational and architectural limitations, we introduce the <u>T</u>emporal <u>E</u>ncoder with <u>L</u>ate <u>F</u>usion (TELF), a lightweight end-to-end predictive model designed to encode the temporal structure of medical code sequences without requiring resource-intensive pre-training. TELF employs an encoder-only architecture that learns medical code embeddings on-the-fly directly from the target cohort, eliminating the need for large-scale pre-training. As a result, TELF enables transformer-based temporal modeling to be performed on standard computing hardware and is readily applicable to both EHR and claims data. Through its late-fusion design, TELF preserves the interpretability of the temporal attention mechanism. In this study, we leverage the enrollment clarity provided by a one-year continuous baseline in an administrative claims database to evaluate TELF across three distinct clinical conditions.

## METHODS

### Data source and study design

To evaluate TELF and compare its predictive performance against established machine learning baselines, we constructed three retrospective cohorts (pancreatic cancer, type 2 diabetes, and heart failure) using longitudinal real-world data from the Optum’s de-identified Clinformatics® Data Mart Database (Optum® CDM). Optum® CDM is derived from a database of administrative health claims for members of large commercial and Medicare Advantage health plans. Clinformatics® utilizes medical and pharmacy claims to derive patient-level enrollment information, health care costs, and resource utilization information. The population is geographically diverse, spanning all 50 US states and is de-identified under the Expert Determination method consistent with Health Insurance Portability and Accountability Act (HIPAA) and Optum® customer data use agreements.

For each of the three conditions, patients were included in the positive class if they received an incident diagnosis for the condition of interest, defined by the presence of specific International Classification of Diseases (ICD) codes, during the identification period (Table 2). The date of the first diagnosis was considered the index date. To ensure complete capture of the medical history (the predictive code sequence), we required continuous enrollment of at least one year prior to the index date (the baseline period). Patients with cohort-specific exclusion ICD codes (Table 2) prior to their index date were excluded to ensure true incident prediction (Figure 1). To align with our objective of developing a resource-efficient framework, we limited the final analytical datasets to computationally accessible sizes. Because type 2 diabetes and heart failure are highly prevalent in the database, we randomly undersampled their positive classes (retaining 10% and 20%, respectively) to approximate the size of the less prevalent pancreatic cancer population.

**Figure 1.**
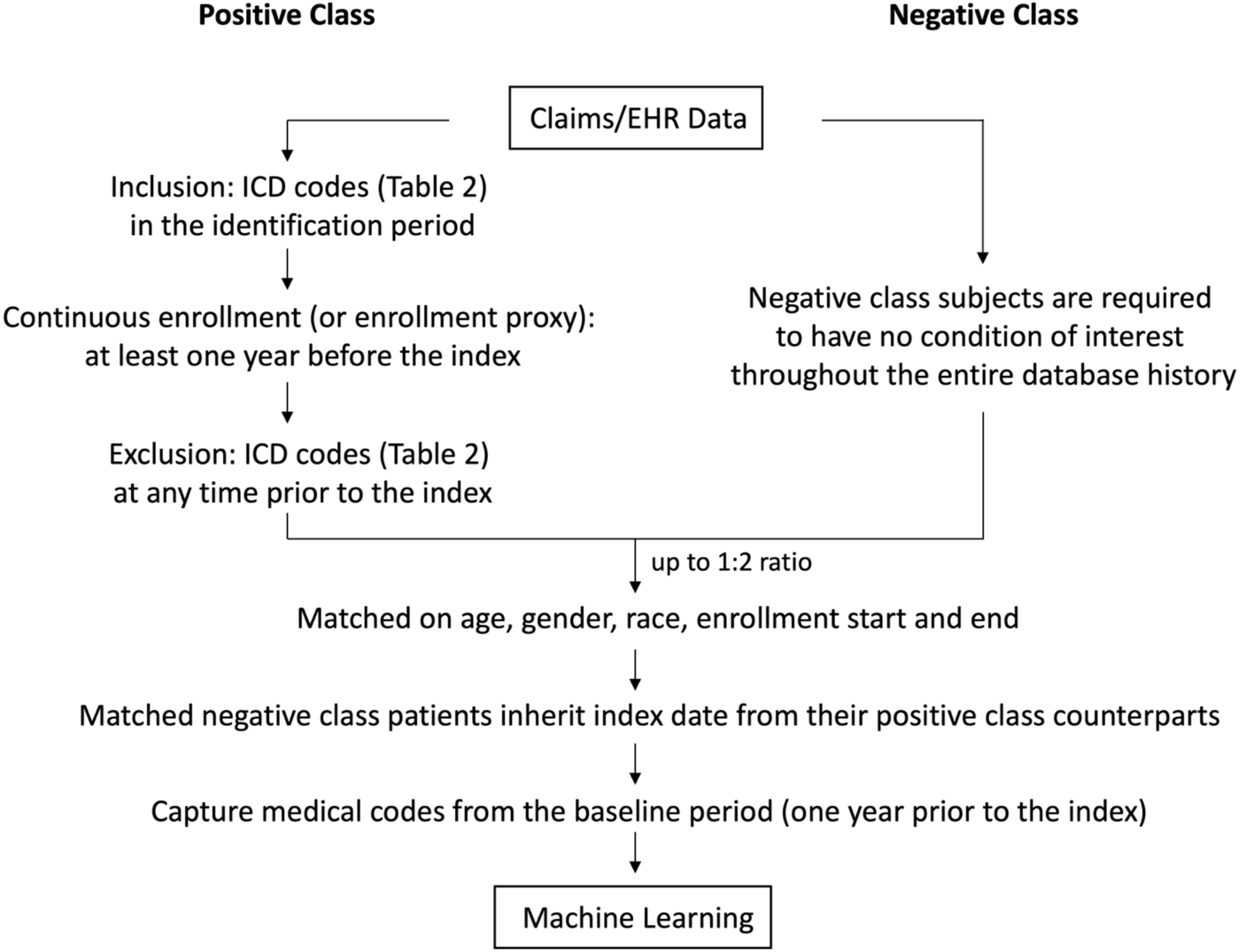
Study design and cohort selection workflow. A systematic flowchart detailing the identification and selection of positive and negative class subjects from longitudinal claims and EHR data. The process includes applying inclusion and exclusion criteria within specific identification periods, enforcing a minimum of one-year continuous enrollment prior to the index date, and implementing a matching algorithm in a ratio of up to 1:2 on age, gender, race, and enrollment start and end. Matched negative class patients inherit index date from their positive class counterparts. The final cohorts undergo temporal feature extraction from the baseline period (one year prior to the index).

**Table 2.** Inclusion and exclusion criteria for cohort definition.

|  | Pancreatic Cancer | Type 2 Diabetes | Heart Failure |
| --- | --- | --- | --- |
| Identification Period | 2016-10-01 ~ 2025-09-30 | 2017-01-01 ~ 2024-12-31 | 2017-01-01 ~ 2024-12-31 |
| ICD for Inclusion | C25.x, 157.x | E11.9, 250.00 | I50.x, 428.x |
| Index Date | First diagnosis in the identification period | First diagnosis in the identification period | First diagnosis in the identification period |
| Continuous Enrollment Requirement | At least one year before the index | At least one year before the index | At least one year before the index |
| ICD for Pre-index Exclusion | C00.x-C96.x, 140.x-209.x | E08.x, E09.x, E10.x, E11.x, E13.x, 249.x, 250.x | I50.x, 428.x |

ICD: International Classification of Disease

To avoid extremely imbalanced analytical datasets, we constructed a matched negative class. The negative class was required to have no record of the condition of interest throughout the entire database history. We matched the negative class patients to the positive class in a ratio of up to 1:2 on age, gender, race, and enrollment start and end. This matching step was implemented to allow the matched negative class patients to inherit a proxy index date from their positive class counterparts. Finally, for both classes, all medical codes that occurred during the one-year baseline period prior to the index date were captured for the machine learning models (Figure 1).

### Data preprocessing

To achieve an optimal level of code granularity while mitigating feature sparsity, all raw ICD diagnosis codes were truncated to the three-character category level. Univariate feature selection was subsequently performed to eliminate weak predictors. The data were flattened into a patient-level binary matrix, and the Phi coefficient calculated between the presence of each medical code and the disease outcome. Codes were deleted if the absolute value of their Phi coefficient was below 0.03. This filtering step efficiently reduced noise and defined the final vocabulary sizes for each condition (ranging from 338 to 505 unique codes, Table 3). Combined with demographic predictors, this patient-level binary matrix served as the feature input for the non-sequential baseline models (e.g., LightGBM, XGBoost, and logistic regression).

**Table 3.** Descriptive analysis of the cohorts.

|  | Pancreatic<br>Cancer | Type 2 Diabetes | Heart Failure |
| --- | --- | --- | --- |
| Cohort size (n) | 53,661 | 78,756 | 72,540 |
| Percent of patients with the outcome | 42% | 45% | 47% |
| Age on the index date (mean ± std) | 72 ± 10 | 63 ± 14 | 76 ± 9 |
| Gender – male (%) | 47% | 48% | 47% |
| Race |  |  |  |
| White (%) | 68% | 57% | 75% |
| Black (%) | 13% | 12% | 12% |
| Asian (%) | 3% | 5% | 2% |
| Other (%) | 16% | 26% | 11% |
| Average number of unique codes | 17 | 14 | 20 |
| Average length of code sequence | 48 | 38 | 58 |
| Vocabulary size | 338 | 442 | 505 |

For the TELF architecture, the temporal integrity of the raw data was preserved. The longitudinal records were filtered to include only the selected vocabulary and chronologically sorted. The data were subsequently transformed to patient-level sequences containing both the ordered medical codes and their corresponding temporal intervals (measured as days prior to the index date). To standardize the input dimensions for the tensor operations, we defined a maximum sequence length of 100 codes. This threshold successfully captured the complete baseline medical history for approximately 90% of the study populations. Sequences exceeding this limit were truncated at the 100^th^ code, while shorter sequences were right-padded with a dedicated <pad> token. Finally, baseline demographic variables were preprocessed: continuous variables z-scored; categorical variables one-hot encoded.

### TELF Model architecture

The TELF architecture consists of three primary components: a temporal sequence encoder, a demographics transformation block, and a late fusion multilayer perceptron (MLP) classifier.

TELF initializes and learns medical code embeddings on-the-fly. For a given patient, the sequence of medical codes is projected into a vector space via a learnable code embedding layer. Simultaneously, the corresponding chronological intervals (days prior to the index date) are passed through a temporal embedding layer. These two representations are added element-wise and subsequently processed by a stack of encoder blocks to accurately represent the patient’s baseline history. Each encoder block utilizes multi-head self-attention to capture long-range dependencies between medical events, followed by a position-wise feed-forward network and standard add-and-norm operations.

To avoid prematurely confounding the self-attention mechanism with non-sequential demographic data, TELF employs a late fusion strategy. Concurrently with the sequence encoding, baseline demographic variables are processed through an independent transformation block. Continuous variables are standardized via z-score normalization, and categorical variables are one-hot encoded. Finally, the high-dimensional temporal output from the last encoder block is pooled and concatenated with the transformed demographic features. This fused feature vector is fed into an MLP classifier to make the final disease risk prediction. In this design, static demographic variables act as post-attention modifiers, preserving the interpretability of the temporal code sequences.

### Training and evaluation

Prior to model training, a sample of 10,000 patients from each disease cohort was reserved as an independent holdout test set to ensure unbiased final evaluation. The remaining data were further divided into an 80% training set and a 20% validation set. TELF was trained using the binary cross-entropy loss function and the AdamW optimizer. To prevent overfitting and optimize generalization, early stopping was implemented, terminating training and restoring the best model weights if the validation loss failed to improve for three consecutive epochs. Final model performance was assessed on the isolated holdout set using the area under the receiver operating characteristic curve (AUC), accuracy, precision, recall, F1 score, and the Matthews correlation coefficient (MCC).

### Attention-based motif mining

To illustrate the interpretability of TELF’s attention mechanism and map the prominent clinical trajectories preceding disease onset, we extracted the self-attention weights from the final encoder layer of the trained TELF model. The attention matrices were averaged across all attention heads to obtain a unified temporal attention map for each patient. To ensure the analysis captured robust predictive signals, we restricted this evaluation to high-confidence true positive patients (high-confidence defined as predicted probability > 0.90).

For each of these high-confidence true positive patients, we calculated the total attention weight received by each medical code across their specific temporal sequence. These patient-level weights were aggregated to identify the four codes that received the highest attention globally. The cohort was further subset to include those patients whose histories contained all four high-attention events. Finally, the three codes occurring most proximally to the disease index date were identified. Sankey plots were generated from these pre-onset sequences to visualize the prominent patient journey motifs leading to the disease outcome.

### Other machine learning models

TELF was evaluated against four benchmark machine learning classifiers: LightGBM, XGBoost, logistic regression, and multilayer perceptron (MLP). As detailed in the Data Preprocessing section, these baseline models utilized the patient-level binary feature matrix supplemented with demographic variables. Logistic regression was implemented in its standard form, as well as with L1 and L2 regularization penalties, using the scikit-learn library. LightGBM and XGBoost were implemented using the lightgbm and xgboost packages, respectively, with hyperparameters tuned via grid search. MLP was implemented using TensorFlow. The network architecture featured six progressively narrowing dense hidden layers with ReLU activations, and a 20% dropout after each hidden layer to mitigate overfitting. The MLP model was trained using the Adam optimizer with a binary cross-entropy loss function. All benchmark models were developed in Python 3.11 and evaluated on the independent holdout test sets.

### Implementation details

All model training, validation, and final evaluations were conducted using the TELF-medium configuration (Table 5). The TELF-medium architecture comprises four attention layers (L = 4), eight attention heads (A = 8), and a hidden embedding dimension of 128 (H = 128). For the processing of longitudinal clinical events, we set the maximum sequence length as 100 codes (tokens). Model optimization was performed using the AdamW optimizer. The primary training hyperparameters included a learning rate of 5e–5, a batch size of 100, and a dropout rate of 0.1.

To demonstrate our goal of democratizing advanced temporal modeling for grassroots clinical researchers, we conducted all training and evaluation exclusively on standard consumer-grade hardware (an Apple MacBook equipped with an M2 processor and 8 GB of memory).

## RESULTS

### Study population characteristics

After applying the inclusion and exclusion criteria and the matching algorithm (Figure 1 and Table 2), a total of 53,661, 78,756, and 72,540 patients were included in the pancreatic cancer, type 2 diabetes, and heart failure analytical cohorts, respectively. With a matching ratio of up to 1:2, the positive class prevalence was successfully balanced, ranging from 42% to 47% across the three datasets. The mean age at the index date was highest in the heart failure cohort (76 ± 9 years) and lowest in the type 2 diabetes cohort (63 ± 14 years). Demographically, the cohorts were predominately White (ranging from 57% to 75%), with varying representation of Black, Asian, and other racial groups. Gender distribution remained relatively even across all three populations (Table 3). The baseline medical code sequences varied in clinical complexity; heart failure presented the most complex histories with an average sequence length of 58 codes drawn from a vocabulary of 505 unique codes, whereas pancreatic cancer and type 2 diabetes exhibited shorter average sequence length of 48 and 38 codes, respectively (Table 3).

### Predictive performance

Evaluated strictly on an independent 10,000-patient holdout test set, TELF (Figure 2) consistently outperformed all baseline machine learning models across the three disease conditions. For pancreatic cancer, TELF achieved the highest predictive performance with an AUC of 0.9150, an F1 score of 0.8374, and a Matthews correlation coefficient (MCC) of 0.7489. The closest baseline models were the tree-based competitors: LightGBM achieved an AUC of 0.9069, an F1 score of 0.8015, and an MCC of 0.6910, while XGBoost achieved an AUC of 0.9044, an F1 score of 0.8017, and an MCC of 0.6903 (Table 4).

**Figure 2.**
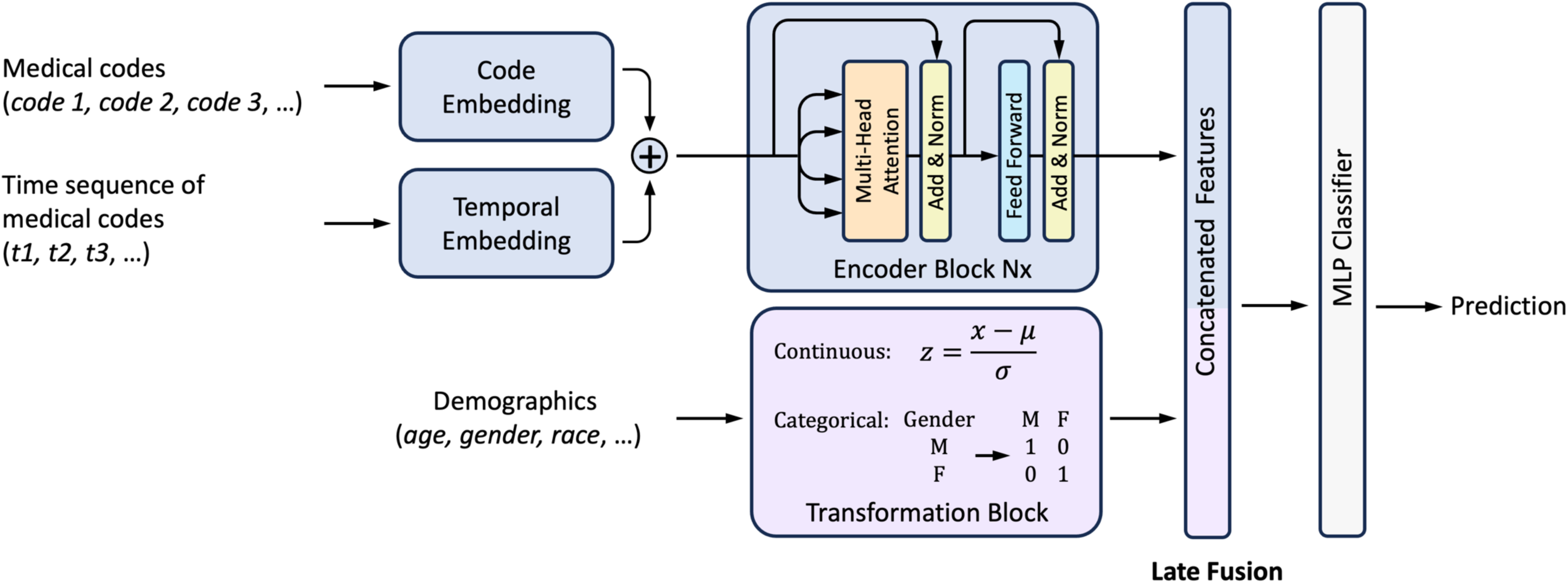
TELF architecture. A detailed diagram of the end-to-end TELF model pipeline. The sequences of medical codes (diagnosis, medication, procedure) are projected into a vector space via a code embedding layer. The corresponding chronological sequences, recorded as days prior to the index date, are passed through a separate temporal embedding layer. These sequences are subsequently processed by multi-head self-attention encoder blocks to capture complex clinical dependencies. Static baseline demographic variables are transformed and concatenated with the pooled temporal output from the last encoder block through a late fusion step for the final disease risk classification.

**Table 4.** Evaluation of TELF and other machine learning models.

| Condition | Model | Accuracy | Precision | Recall | F1 | MCC | AUC |
| --- | --- | --- | --- | --- | --- | --- | --- |
| Pancreatic Cancer | TELF | 0.8761 | 0.9317 | 0.7605 | 0.8374 | 0.7489 | 0.9150 |
|  | XGBoost | 0.8487 | 0.8905 | 0.7290 | 0.8017 | 0.6903 | 0.9044 |
|  | LightGBM | 0.8489 | 0.8929 | 0.7271 | 0.8015 | 0.6910 | 0.9069 |
|  | LR | 0.8460 | 0.8969 | 0.7152 | 0.7958 | 0.6858 | 0.9014 |
|  | LR with L1 | 0.8463 | 0.8994 | 0.7135 | 0.7957 | 0.6868 | 0.9017 |
|  | LR with L2 | 0.8454 | 0.8981 | 0.7123 | 0.7945 | 0.6849 | 0.9009 |
|  | MLP | 0.8448 | 0.8794 | 0.7302 | 0.7979 | 0.6814 | 0.9038 |
| Type 2 Diabetes | TELF | 0.7458 | 0.7444 | 0.6717 | 0.7062 | 0.4852 | 0.8199 |
|  | XGBoost | 0.7277 | 0.7389 | 0.6205 | 0.6746 | 0.4485 | 0.7908 |
|  | LightGBM | 0.7251 | 0.7333 | 0.6216 | 0.6729 | 0.4430 | 0.7916 |
|  | LR | 0.7235 | 0.7402 | 0.6040 | 0.6652 | 0.4403 | 0.7800 |
|  | LR with L1 | 0.7239 | 0.7410 | 0.6040 | 0.6655 | 0.4412 | 0.7803 |
|  | LR with L2 | 0.7231 | 0.7400 | 0.6031 | 0.6646 | 0.4395 | 0.7802 |
|  | MLP | 0.7297 | 0.7243 | 0.6550 | 0.6879 | 0.4524 | 0.7871 |
| Heart Failure | TELF | 0.7983 | 0.8061 | 0.7566 | 0.7806 | 0.5953 | 0.8721 |
|  | XGBoost | 0.7767 | 0.7850 | 0.7286 | 0.7558 | 0.5519 | 0.8535 |
|  | LightGBM | 0.7733 | 0.7725 | 0.7398 | 0.7558 | 0.5449 | 0.8546 |
|  | LR | 0.7718 | 0.7964 | 0.6970 | 0.7434 | 0.5435 | 0.8466 |
|  | LR with L1 | 0.7712 | 0.7965 | 0.6951 | 0.7423 | 0.5423 | 0.8469 |
|  | LR with L2 | 0.7717 | 0.7968 | 0.6961 | 0.7431 | 0.5433 | 0.8466 |
|  | MLP | 0.7737 | 0.7712 | 0.7434 | 0.7570 | 0.5457 | 0.8503 |

TELF: Temporal Encoder with Late Fusion
LR: Logistic Regression
MLP: Multilayer Perceptron
MCC: Matthews Correlation Coefficient
AUC: Area under the receiver operating characteristic curve

This superiority in disease risk prediction was maintained in the type 2 diabetes and heart failure cohorts, where TELF showed AUCs of 0.8199 and 0.8721, respectively (Table 4). Across all three conditions, the tree-based models (XGBoost and LightGBM) generally outperformed logistic regression and standard multilayer perceptron (MLP), but failed to match the temporal pattern recognition capabilities of the TELF architecture (Table 4). The receiver operating characteristic (ROC) curves confirmed TELF’s superior sensitivity and specificity trade-off compared with other ML models (Figure 3).

**Figure 3.**
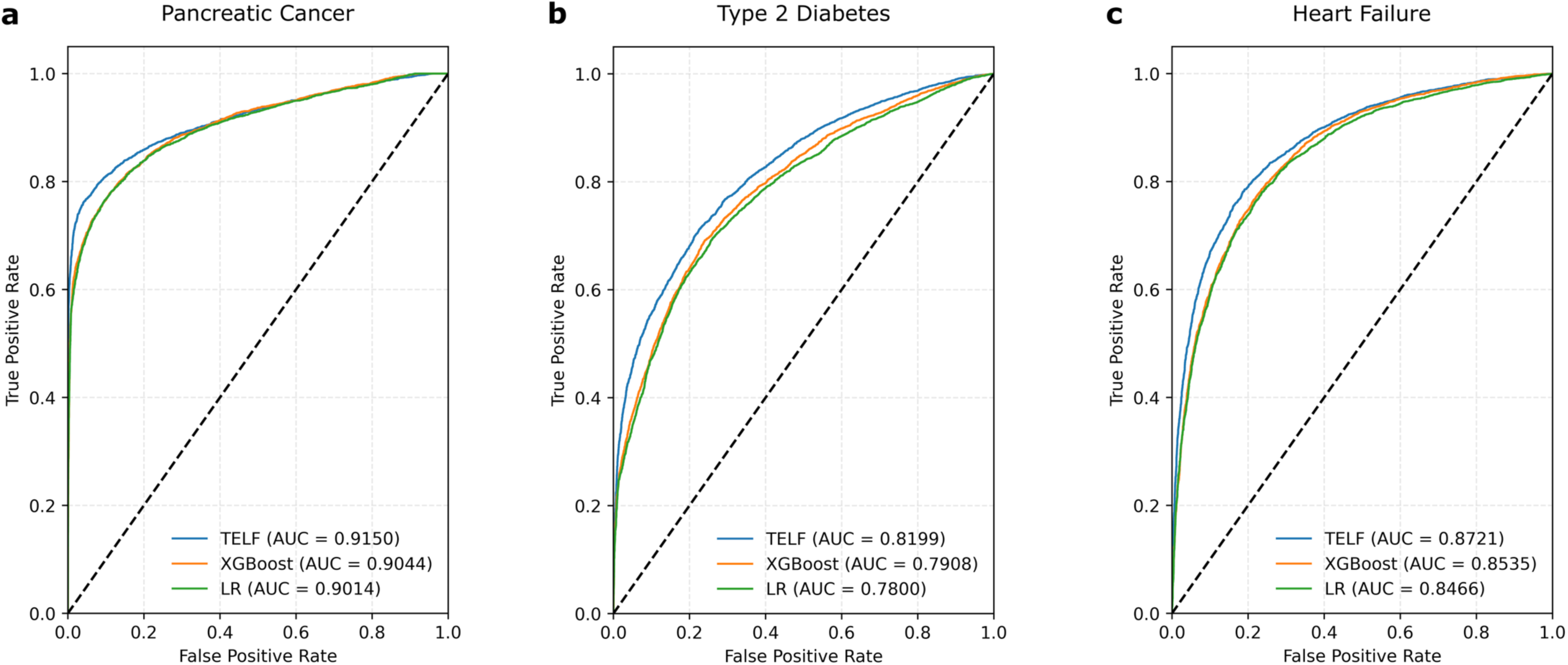
Receiver Operating Characteristic (ROC) curves for disease prediction. Comparative ROC curves are shown for TELF (blue), XGBoost (orange), and Logistic Regression (green) for three distinct conditions: (a) pancreatic cancer, (b) type 2 diabetes, and (c) heart failure. Performance was strictly evaluated on an independent holdout test sets for each cohort.

### Attention-based temporal motifs

Beyond improved prediction accuracy, the TELF architecture demonstrated clinical interpretability by identifying specific temporal sequences prior to the disease onset. By extracting and aggregating the self-attention weights from high-confidence true positive patients, the medical codes that received the highest global attention were identified and distinct patient journey motifs were mapped. For example, the Sankey diagram in Figure 4 illustrates the prominent progression of clinical events leading to the disease onset in the pancreatic cancer cohort. The visualization maps the temporal transitions between the highest-attended pre-onset code categories, including other diseases of the pancreas (K86), abdominal and pelvic pain (R10), unspecified jaundice (R17), and symptoms concerning food/fluid intake (R63). This motif mining demonstrates TELF’s capacity to learn and identify interpretable, attention-based patient journey summaries. By extracting distinct pre-onset trajectories, such as the specific temporality of unspecified jaundice and abdominal pain, TELF provides clinicians with valuable epidemiological insights that traditional machine learning models cannot offer.

**Figure 4.**
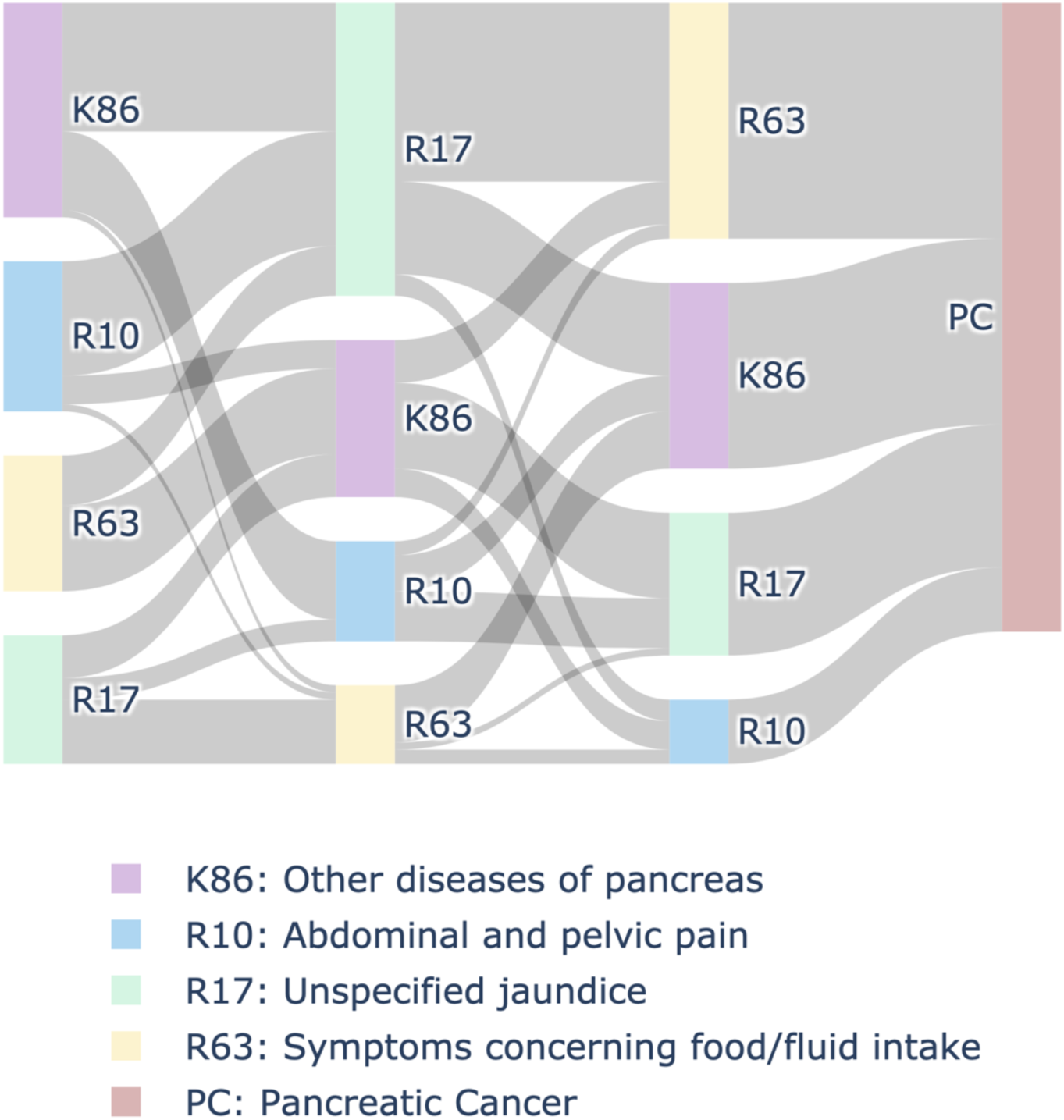
Attention-based patient journey motifs. A Sankey diagram visualizing the prevalent trajectories leading to disease onset in the pancreatic cancer cohort. The visualization aggregates temporal clinical sequences from high-confidence (predicted probability > 0.90) true positive patients by leveraging self-attention weights. The diagram illustrates the temporal sequence and flows between the four diagnostic codes that received the highest attention: other diseases of pancreas (K86), abdominal/pelvic pain (R10), unspecified jaundice (R17), and symptoms concerning food/fluid intake (R63).

## DISCUSSION

In this study, we developed and validated the <u>T</u>emporal <u>E</u>ncoder with <u>L</u>ate <u>F</u>usion (TELF), a lightweight end-to-end predictive deep learning model designed for longitudinal administrative claims and EHR data. TELF learns medical code embeddings on-the-fly, thereby bypassing the resource-intensive pre-training step characteristic of many existing sequence models. Evaluated across three distinct chronic conditions, pancreatic cancer, type 2 diabetes and heart failure, TELF consistently outperformed machine learning benchmark models on an independent holdout test set. Beyond achieving superior predictive accuracy, TELF preserved the interpretability of its self-attention mechanism, enabling the identification of key patterns in the pre-onset patient journey that can inform public health and clinical decision-making directly.

The current AI landscape is largely defined by a hardware arms race - one in which access to specialized compute infrastructure has become the primary competitive moat [16]. Driven by major technology corporations, the rapid deployment of large language models (LLMs) has established a paradigm that relies heavily on immense computational resources, dedicated cooling infrastructure, and massive carbon footprints [16, 17]. Following this trend, clinical informatics has largely adopted a resource-intensive, two-stage approach: massive foundation models are first pre-trained on large-scale healthcare data and subsequently fine-tuned on specific downstream tasks [7]. This has created a misconception that rigorous sequence modeling in healthcare requires large corpora of training data and cost-prohibitive GPU clusters, rendering temporality-based predictive clinical research a luxury accessible only to well-resourced institutions. While conceptually impressive, current foundation models offer minimal practical benefit to grassroots researchers that lack access to proprietary healthcare data and high-performance computing clusters.

TELF was explicitly designed to bypass this computational barrier. By optimizing the architectural hyperparameters for standard cohort sizes (10,000 to 100,000 patients), we developed a highly capable sequence model that scales from roughly 1.2 million to 6.3 million parameters (Table 5). Our primary configuration, TELF-medium, utilizes an embedding dimension of 128, eight attention heads, and four attention layers, resulting in approximately two million trainable parameters. At this scale, TELF demonstrates that accurate sequence-based temporal modeling does not require cost-prohibitive GPUs; indeed, TELF-medium is fully executable on standard consumer hardware, and was trained end-to-end on an Apple M2 laptop with only 8 GB of memory. This computational accessibility ensures that TELF is not merely a theoretical exercise, but a highly practical and deployable framework for hospital data science teams and machine learning engineers that conduct predictive analytics in real-world settings.

**Table 5.** Architectural details of TELF models.

| Model | # Layers | # Attention heads | Hidden size | # Parameters | Recommended hardware |
| --- | --- | --- | --- | --- | --- |
| TELF-base | 2 | 4 | 64 | ~1.2 million | Standard or advanced CPU |
| TELF-medium* | 4 | 8 | 128 | ~2.0 million | Advanced CPU or entry-level GPU |
| TELF-large** | 6 | 8 | 256 | ~6.3 million | Dedicated GPU |

\* Primary configuration for training, validation and evaluations in this study
\*\* Proposed architecture only; not evaluated in this study as the primary study objective is to demonstrate high predictive performance under consumer-grade hardware constraints

In addition to computational accessibility, a more fundamental limitation of the pre-trained foundation models lies in the generalizability of the learned embeddings across different datasets. The assumption that embeddings pre-trained on a single massive corpus can be reliably transferred and fine-tuned for condition-specific predictive tasks on a distinct database is methodologically flawed. This challenge manifests across multiple dimensions. First, a key issue arises from cross-national differences in healthcare-seeking behavior, which fundamentally alter the structure and cadence of EHR and claims data. In the UK, universal coverage under the National Health Service (NHS) lowers the threshold for primary care visits, resulting in more frequent general practitioner (GP) encounters [18], as seen in the Clinical Practice Research Datalink (CPRD) [19] used to train BEHRT [3]. In contrast, most US patients are covered by commercial insurance and therefore seek care more conservatively due to deductibles and co-pays [18]. Consequently, embeddings pre-trained on UK data are poorly suited for transfer to US cohorts and can embed behavioral biases into downstream predictive models. Second, discrepancies in clinical coding ontologies further undermine transferability. For instance, BEHRT was trained on CPRD data that rely on Read codes for primary care and ICD-10 codes for secondary care [19], both of which were mapped to the UK-specific Caliber coding system. Applying these pre-trained embeddings to standard US claims or EHR data (which primarily use ICD codes for diagnosis) requires cumbersome cross-walking that inevitably results in a loss of code granularity. Third, even within a single country, dataset-specific characteristics can compromise the utility of transferred embeddings. For example, ClinicalBERT [4] was trained on the MIMIC-III critical care database [20], in which diagnosis codes are collected from patients in intensive care units (ICU). The learned embeddings inherently encode a skewed distribution of disease severity that is unrepresentative of general patient populations. In addition, privacy-preserving date-shifting in the MIMIC-III data distorts the true chronological spacing of medical events. Consequently, using these ICU-derived embeddings for prediction tasks on standard US claims or EHR data is highly unreliable. In contrast, TELF’s end-to-end design completely circumvents these transferability issues. By learning the semantic meanings and temporal dynamics of medical codes directly from the target dataset, TELF generates embeddings that are intrinsically aligned with the study population’s specific coding standards, healthcare-seeking behaviors, and chronological rhythms.

A core driver of TELF’s efficiency and interpretability is its late-fusion architecture. Recent sequence models in healthcare, such as BEHRT [3], TransformEHR [8], DuETT [12], Hi-BEHRT [13], and ExBEHRT [14], frequently utilize early or complex multimodal fusion, in which static demographic variables (such as age, sex, or race) are embedded directly alongside sequences of medical codes. This early-fusion approach relies on a strong and counter-intuitive assumption: that the intrinsic clinical meaning of a medical code changes based on a patient’s demographics. In real-world settings, while demographics can influence baseline disease risk, the clinical definition of a medical code remains static. By employing late fusion, TELF first processes the temporal code sequence and reserves demographic predictors as risk modifiers only at the final classification step. This decoupling maintains the absolute integrity of the attention mechanism and prevents chronological signals from being prematurely adjusted by static demographic predictors.

Because the attention mechanism remains unconfounded by demographics, TELF transcends the “black box” limitations of traditional machine learning models by enabling attention-based motif mining. This capability has significant implications for real-world clinical practice. For instance, our motif analysis of the pancreatic cancer cohort identified distinct patient journeys, revealing subsets of patients who experienced unspecified jaundice prior to abdominal or pelvic pain, versus those who manifested pain prior to jaundice (Figure 4). If a clinician or health system observes a sudden localized increase in the prevalence of a specific motif, it could signal an emerging public health priority [21, 22]. Such shifts in symptom presentation can be attributed to underlying dietary changes, localized pollutants affecting the biliary tract, or new occupational exposures [23]. By presenting these aggregated patient journey motifs, TELF equips clinicians and epidemiologists with actionable targets for investigation that the traditional non-sequential models cannot provide.

While TELF excels in disease risk prediction across three distinct cohorts, it is important to note its boundary conditions in epidemiological research. The architecture of TELF is designed to leverage the temporality of clinical events in chronic and progressive conditions. For acute conditions with little or no dependency on long-term clinical history, such as the resolution of jaundice after a hepatitis A virus infection [24, 25], the temporal sequence from the baseline period offers minimal predictive value. In such cases, TELF’s performance would likely converge with, or offer no distinct advantage over, the traditional machine learning models.

Furthermore, the work presented herein highlights several avenues for future architectural refinement. Currently, TELF relies on a standard multilayer perceptron (MLP) for its final classification step. Future iterations could explore replacing this MLP layer with an optimized tree-based classifier, such as XGBoost or Random Forest, to potentially enhance the late fusion of the high-dimensional temporal vector with the static demographic predictors. More importantly, while TELF can efficiently learn the semantic meanings of discrete diagnosis, procedure and medication codes, its current architecture is unable to natively process continuous numeric values from certain laboratory results. The integration of laboratory tests presents a unique architectural challenge; while the vast majority of standard medical codes represent a simple binary presence or absence, many laboratory tests inherently carry numeric magnitudes. In existing literature, transformer-based EHR models frequently circumvent this issue by discretizing or binning continuous lab values into categorical tokens (e.g., “normal,” “abnormal-high”) or quartile ranks to force compatibility with standard discrete embedding layers [26, 27]. Other prominent architectures process lab results indirectly by tokenizing them as text strings extracted from unstructured clinical notes [7]. However, both arbitrary categorization and text-based tokenization are fundamentally information-losing processes, as they diminish the precise magnitude and variance of the raw diagnostic results. Models explicitly designed to process extended clinical histories primarily focus on optimizing the attention mechanisms for long sequences of discrete events, rather than natively solving the continuous value integration problem [13, 14]. Therefore, rather than relying on coarse-grained discretization, a critical next step is the development of a dedicated continuous value projection mechanism. Integrating continuous time-series forecasting layers [28] that can embed raw numeric laboratory values directly into the shared dense vector space alongside the learned discrete code embeddings represents the definitive path forward in capturing a complete multi-modal clinical history.

In conclusion, the <u>T</u>emporal <u>E</u>ncoder with <u>L</u>ate <u>F</u>usion (TELF) represents a highly accessible, end-to-end framework for modeling longitudinal claims and EHR data. By learning medical code embeddings directly from the target cohort and decoupling static demographic variables from chronological medical events, TELF achieves superior predictive performance across diverse conditions without requiring the prohibitive computational overhead of massive pre-trained foundation models. Importantly, TELF transcends mere classification by leveraging its native self-attention mechanism to facilitate the mining of patient journey motifs preceding disease onset. This synthesis of predictive accuracy, computational efficiency, and attention-driven motif mining provides health systems, clinicians, and epidemiologists with a powerful yet accessible tool not only for forecasting disease risk, but also for characterizing the evolving clinical trajectories of specific patient populations.

## Declaration statements

### Data availability statement

The datasets generated or analyzed during the current study are not publicly available due to confidentiality constraints and Optum® customer data use agreements. Optum’s de-identified Clinformatics® Data Mart (CDM) Database is proprietary to Optum.

### Code Availability

The codes for data pre-processing, TELF model architecture, late fusion, and model training and evaluation are publicly available at https://github.com/unclevanya1899/TELF/.

## Acknowledgements

This work was sponsored by Gilead Sciences, Inc. The authors would like to thank Robert Stafford, Michael Crowley and Amanda M Kelly for their assistance with the description of Optum® CDM data.

## Author contributions

LL and ZZ designed the study. LL analyzed the data. Both authors interpreted the data. LL drafted the manuscript, and ZZ revised it. Both authors approved the final version of the manuscript.

